# Effectiveness of integrating cervical cancer prevention strategies into HIV care programmes: A mixed-methods systematic review protocol

**DOI:** 10.1101/2024.07.07.24309237

**Authors:** Kimeshnee Govindsamy, Susanne Noll, Ntombifuthi Blose, Edina Amponsah-Dacosta

**Affiliations:** Division of Epidemiology and Biostatistics, School of Public Health and Family Medicine, University of Cape Town, Western Cape, South Africa; Vaccines for Africa Initiative (VACFA), Faculty of Health Sciences, School of Public Health and Family Medicine, University of Cape Town, Cape Town, South Africa; Health Systems Trust, Durban, South Africa

## Abstract

**Introduction:** Cervical cancer, which is the fourth most frequently diagnosed cancer among women globally, remains a significant health burden despite being preventable and treatable, exposing gaps in accessing prevention and control services. Adolescent girls and young women (AGYW) face heightened risk of persistent HPV infection, a primary cause of cervical cancer. The overlap of cervical cancer and HIV exacerbates public health challenges, especially in resource-limited areas, urging intensified efforts in bolstering prevention and control measures. Integrating HPV vaccination, cervical cancer screening, treatment of precancerous lesions and educational interventions into HIV care programs shows promise in effectively addressing this dual burden.

**Methods:** To evaluate the effectiveness of integrating cervical cancer prevention strategies within HIV care programs, a mixed-methods systematic review will be conducted. A comprehensive Boolean search for literature published between 2006 to present and indexed in PubMed, Cochrane Library, EBSCO Host, Web of Science, Scopus, and Google Scholar will be conducted, without imposing any language restrictions. This review will be conducted in alignment with the Joanna Briggs guidelines on systematic reviews together with the Preferred Reporting Items for Systematic Reviews and Meta-Analyses (PRISMA) guidelines. Data from eligible studies will be extracted and synthesized, and their quality assessed.

**Discussion:** There is limited understanding of the effectiveness of integrating cervical cancer prevention and HIV care in the real-world setting. While some studies touch on integration, focus tends to be on cervical cancer screening alone, neglecting vaccination, treatment of precancerous lesions, and education programs. Previous reviews on this focus are outdated, surpassing six years. This systematic review aims to fill these evidence gaps by thoroughly evaluating the challenges and opportunities associated with integrating the full complement of HPV prevention strategies and HIV care programs. The anticipated findings could enhance service delivery models aimed at reducing cervical cancer incidence and mortality among AGYW living with HIV.

**Trial registration:** Systematic review registration: PROSPERO registration number: CRD42024535821.

## Introduction

### Background

Cervical cancer ranks as the fourth most frequently diagnosed cancer among women globally, with current global estimates from 2022 indicating that 660 000 women are diagnosed with cervical cancer and 350 000 women die from this disease annually, according to the World Health Organization (WHO) [1]. Persistent infection with the Human Papillomavirus (HPV) following sexual transmission is the primary sufficient cause of cervical cancer [2]. Furthermore, cervical cancer is the most common HPV-related disease [2]. Most HPV infections resolve spontaneously and do not cause symptomatic disease. However, persistent infection with specific high-risk HPV types (most frequently HPV types 16 and 18) may lead to pre-cancerous lesions and if untreated these lesions may progress to invasive cervical cancer [2]. According to Okunade [2] approximately 99.7% of cervical cancer cases are caused by persistent high-risk HPV infection. HPV is estimated to infect approximately 291 million women globally, with a significantly higher prevalence among women under the age of 25 years [3].

There are significant disparities in the prevalence of cervical cancer globally which highlights variations in accessibility, coverage, and quality of preventative and control measures as well as the prevalence of risk factors for cervical cancer. More than 85% of cervical cancer cases and deaths occur in low- and middle-income countries (LMICs) where these measures are suboptimal whilst the prevalence of risk factors are usually higher [4]. This inequality gap continues to widen, as substantial declines in cervical cancer incidence rates have been observed in high-income countries (HICs) with some nations even forecasted to be progressing toward the goal of cervical cancer elimination in the coming decades [5]. This can be attributed to the effectiveness of routine cervical cancer prevention programs in these regions. In contrast, in certain sub-Saharan African regions and in several European and western Asian countries, rates have either increased or remained relatively stable at elevated levels [5]. This is attributed to the suboptimal provision and utilization of prevention programs in these settings.

Although HPV is the underlying sufficient cause of cervical cancer there are other risk factors associated with this disease which include smoking, increased parity, long-term use of oral contraceptives as well as infection with human immunodeficiency virus (HIV) [2]. Studies have shown that women who are HIV-positive face a six-fold increased risk of developing cervical cancer compared to those without HIV, which further exacerbates the burden of cervical cancer [5]. This heightened risk stems from a multifaceted interplay of biological and societal factors. Among these are the direct impact of HIV on the immune regulation of HPV, accelerated disease advancement in HIV-positive women, extended life expectancy due to antiretroviral therapy, and obstacles such as stigma, poverty, and gender-related barriers that hinder women from accessing timely care [4]. Compelling evidence indicates that women infected with HIV face an elevated risk of persistent infection with multiple types of HPV at an early age, specifically between the ages of 13 and 18 years, which contributes to an increased likelihood of developing cervical cancer at a younger age [3]. This dual burden of HPV and HIV poses a complex challenge to healthcare systems worldwide, especially among adolescent girls and young women (AGYW) living with HIV.

Cervical cancer can be prevented, treated, and ultimately eliminated as a public health concern through primary, secondary, and tertiary prevention measures. These prevention strategies differ among females living without and with HIV as outlined in Fig 1, as per the WHO guidelines [6]. Primary prevention measures include HPV vaccination and educational interventions that create awareness related to cervical cancer risks and prevention and control strategies. Secondary prevention includes cervical cancer screening and treatment of precancerous lesions. Tertiary prevention includes the treatment of invasive cancer.

**Fig 1.**
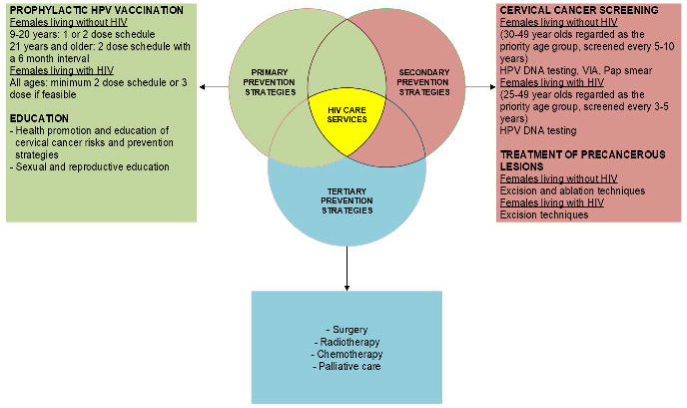
Summary of cervical cancer prevention strategies as per WHO guidelines [6].

### Primary prevention

Prophylactic HPV vaccination has been regarded as the most effective long-term strategy for preventing HPV infection, cervical cancer and other cancers associated with HPV [4]. HPV vaccines are most effective if administered prior to HPV-exposure and therefore is primarily targeted to adolescent girls (9-14 years of age), prior to sexual debut [7]. The first HPV vaccine was licensed in 2006 and currently there are six licensed prophylactic vaccines available, all of which are recommended for global use by the WHO [7]. These include three bivalent vaccines, two quadrivalent vaccines and one nonvalent vaccine [4]. HPV vaccines are designed to prevent over 95% of HPV infections caused by the common high-risk HPV types 16 and 18 as well as some cross protection against other less common HPV types that can cause cervical cancer [1]. Vaccination is a powerful preventative strategy as it targets the underlying cause and moreover widespread vaccination creates herd immunity, benefiting even those who cannot receive vaccines. According to WHO recommendations, females aged 9-20 years should receive one- or two-dose schedules and females 21 years and older should receive two-doses with a six- month interval [7]. Females living with HIV at all ages should receive three doses (or two doses if feasible) [7]. The high prevalence of HPV infections in HIV positive females emphasizes how important it is to vaccinate this population. HPV vaccines are a safe and effective tool that when combined with other cervical cancer prevention strategies can significantly reduce the incidence of cervical cancer and its associated mortality.

Educational interventions encompass health education initiatives that are designed to enhance an individual’s understanding and awareness of health-related matters, in this instance matters relating to cervical cancer prevention strategies, which ultimately would lead to a positive change in their behaviour [8]. These interventions play a crucial role in creating awareness and imparting knowledge on the importance of cervical cancer prevention strategies and risks. In addition, sexual and reproductive education, tailored to age and culture, that promotes safer sexual practices also aids in creating awareness and potential behavioural changes. In providing the necessary information females are empowered to make informed decisions about their health, seek HPV vaccination services and timely screening, as well as adopt proactive measures and ultimately reduce their risk of developing cervical cancer.

### Secondary prevention

To prevent cervical cancer females can undergo various cervical screening tests to detect precancerous cells and thereafter receive the appropriate pre-treatment to reduce the risk of progression to invasive cervical cancer. The conventional approach for screening involves cytology screening (conventional or liquid based), commonly known as Papanicoloau (Pap) test or pap smear [9]. Positive cytology results lead to confirmation through colposcopy, and subsequent biopsies of suspicious lesions for histologically confirmed diagnosis [9]. Over the past 15 years, additional screening strategies have been introduced including visual inspection with acetic acid (VIA) and molecular tests such as high-risk HPV DNA testing [9]. In 2021 the WHO recommended HPV-DNA detection as the primary screening method particularly for females living with HIV [6]. The WHO identifies females living without HIV in the age group 30-49 years as a priority for screening and should be screened every five to ten years with a pap smear, VIA technique or HPV-DNA testing [6]. For females living with HIV, those aged 25-49 years are identified as a priority group and should be screened every three to five years preferably with HPV-DNA testing or other screening tests as available [6]. After the age of 50 years the WHO suggests screening is stopped after two consecutive negative screening results in females living with and without HIV [6]. More recently newer techniques have been developed: other molecular tests such as those relying on HPV messenger ribonucleic acid (mRNA), oncoprotein detection or DNA methylation; more objective assessments conducted on cytological samples such as p16/Ki67 dual staining; and enhanced visual inspection tests utilizing artificial intelligence/ machine learning platforms such as automated visual evaluation of digital images [6].

In resource-limited settings, the predominant approach for treatment of cervical abnormalities is tissue ablation techniques such as cryotherapy or thermal ablation [10]. Ideally, if the patient qualifies for ablative treatment, it should be performed immediately during the same visit as the positive screening test (single-visit approach). However, at times this may not be feasible in some healthcare facilities and therefore a second visit is required (multiple-visit approach). For women who are not eligible for ablation or in HICs the main approach for treatment involves the excision of histologically confirmed cervical abnormalities [10]. Excisional treatment options include loop electrosurgical excision (LEEP), large loop excision of the transformation zone (Lletz) or cone excision [10].

The WHO recommends two approaches to screening and treatment namely, the “screen-and-treat” and the “screen, triage and treat” approach [6]. In the “screen-and-treat” approach treatment is administered to females solely based on a positive primary screening test (which means there is no secondary screening test and no histopathological diagnosis) [6]. In the “screen, triage and treat” approach the treatment decision is based on a positive result from the initial screening test, followed by a positive outcome in a subsequent test (referred to as a “triage” test), with or without histologically confirmed diagnosis [6]. For females in the general population, the WHO recommends HPV DNA detection as the screening method in a “screen-and-treat” or “screen, triage and treat” approach starting at the age of 30 years with regular screening every five to ten years [6]. For females living with HIV, it is recommended that HPV DNA detection be used in a “screen, triage and treat” approach only starting at the age of 25 years with regular screening every three to five years [6].

Despite the development and implementation of cervical cancer prevention strategies this disease continues to be a public health concern, particularly in LMICs. As a result of this in 2020 the WHO introduced a “global strategy” to accelerate the elimination of cervical cancer [6]. This strategy proposes the following targets: 90% of adolescent girls globally should be vaccinated against HPV; 70% of women should undergo HPV screening and 90% of women diagnosed with cervical cancer should receive suitable follow-up treatment [6]. The aim of this global strategy is to reduce the incidence of cervical cancer to below a threshold of 4 cases per 100 000 women-years in every country [11]. Meeting these objectives set out by the WHO requires reconsideration of existing strategies to accelerate the adoption of HPV vaccination, cervical cancer screening and timely treatment of precancerous lesions, whilst optimizing rational resource allocation and use. The WHO advocates for the integration of these prevention strategies in other healthcare systems to further enhance its effectiveness [12].

There has been a growing interest in integrating cervical cancer prevention strategies into HIV care programs [13]. Such integration can potentially improve access to cervical cancer screening, early detection, and pre-treatment services for AGYW living with HIV. Additionally, it may offer opportunities for the efficient utilization of established healthcare resources and strengthen the health system’s capacity to address both HIV and cervical cancer. Integrated healthcare approaches aim to leverage existing HIV care infrastructure to provide cervical cancer preventative services, which can encompass the full complement of prevention strategies. The integration not only streamlines healthcare delivery but also capitalizes on the regular contact that AGYW living with HIV have with healthcare providers. Supporting evidence highlighting the feasibility and outcomes of such integration programs has been published. For instance, Mwanahamuntu *et al*., [14] conducted a study on the integration of cervical cancer prevention services into HIV care services in Zambia and reported that over the course of 2.5 years more than 20 000 women had undergone cervical cancer screening following integration. Furthermore, studies conducted in Kenya, Mozambique and Botswana have reported cervical cancer screening within HIV care services to be feasible, acceptable, and effective [15–17].

Despite the potential benefits of integrating cervical cancer prevention and HIV care, there is still a limited understanding of its effectiveness in the real-world setting. As such, it is important to systematically assess the effectiveness of integrating such programs across various contexts.

This systematic review seeks to comprehensively assess existing evidence, by employing a mixed methods approach, to provide a holistic understanding of the outcomes and implications of integrating cervical cancer prevention strategies into HIV care programs. This review will delve into the uptake of HPV vaccination, cervical cancer screening, precancerous treatment, and educational interventions aimed at enhancing personal urgency and positive behavioural change. In addition, we will describe the knowledge, awareness, and willingness of AGYW living with HIV with regards to utilizing and adhering to these strategies following integration into existing HIV care programs. Given the increasing global burden of cervical cancer, the continued burden of HIV, and the increased risk of HPV infections in AGYW living with HIV, this review will hold significant relevance and urgency for public health initiatives aimed at improving the health and well-being of AGYW living with HIV as well as reducing the incidence and mortality rates of cervical cancer.

### Rationale for the review

Cervical cancer is the fourth most common cancer among women globally, despite it being a preventable and curable health issue through universal access to effective cervical cancer prevention and control programs. AGYW living with HIV are a population of particular concern as they are at an increased risk of persistent infection with multiple types of HPV. The co-occurrence of cervical cancer and HIV represents a significant public health challenge, particularly in resource-limited settings and requires more effort to be placed on improving current cervical cancer prevention and control measures. Cervical cancer prevention strategies should encompass multidisciplinary approaches that include HPV vaccination, screening, precancerous treatment, and educational interventions. Utilising existing HIV care programmes as a means of integrating cervical cancer prevention strategies into HIV routine care provides an effective way for improving cervical cancer prevention for the most at-risk population and should be prioritised. Existing literature primarily focuses on isolated aspects of either HIV care or cervical cancer prevention, or some but not all preventative strategies, thus leaving a fragmented understanding of the holistic benefits of integrating these public health concerns. Studies have considered integration of these programs [15–17], although, more focus is placed on cervical cancer screening as a prevention strategy as opposed to comprehensive approach that includes vaccination, education, and pre-treatment. Moreover, the search end dates of these reviews are outdated -, being more than six years old- and do not consider recent recommendations and advancements in the field. This review aims to provide a comprehensive assessment of the effectiveness, knowledge, awareness, and willingness among AGYW living with HIV, related to integrating cervical cancer prevention strategies, which includes vaccination, screening, precancerous treatment, and educational interventions, into existing HIV care programs. A mixed methods systematic review is an ideal research design for this study as it allows for an overall examination of both quantitative and qualitative data. The quantitative evidence will provide insights into the effectiveness of the integration strategy, whilst the qualitative evidence can shed light on the knowledge, awareness, and willingness of AGYW living with HIV. Findings from this review will have the potential to reform current policy and practice, thereby contributing to improving health outcomes for AGYW living with HIV and reduce cervical cancer incidence and mortality.

## Research aims and objectives

### Aim

To describe the effectiveness of integrating cervical cancer prevention strategies into existing HIV care programs.

### Objectives

1. To identify and describe integration models for cervical cancer prevention strategies and HIV care programs.
2. To assess the effectiveness of integrating cervical cancer prevention strategies into existing HIV care programs.

2.1 To assess the uptake of HPV vaccination, cervical cancer screening, precancerous treatment, and educational interventions among AGYW living with HIV following integration into existing HIV care programs.
2.2 To assess the knowledge, awareness, and willingness of AGYW living with HIV to utilise and adhere to cervical cancer prevention strategies following integration into existing HIV care programs.

### Methods

A comprehensive mixed methods review that evaluates primary studies employing qualitative, quantitative, and mixed methods will be conducted in alignment with the Joanna Briggs guidelines on mixed methods systematic reviews together with the Preferred Reporting Items for Systematic Reviews and Meta-Analyses (PRISMA) guidelines [18,19]. The protocol for this review was developed in accordance with the PRISMA-Protocol (PRISMA-P) guidelines and checklist (S1 Checklist. PRISMA-P 2015 checklist.) and is registered with the International Prospective Register of Systematic Reviews (PROSPERO) (ID: CRD42024535821), any changes to the published record will be reported [20]. The proposed timeline for this review is February 2024 to February 2025.

### Eligibility Criteria

#### Inclusion Criteria

1. All primary research studies (including those with quantitative, qualitative, and mixed methods research designs) reporting findings on the effectiveness, knowledge, awareness, and willingness related to integrating cervical cancer prevention strategies (vaccination, education, screening, treatment of precancerous lesions, and educational interventions) into existing HIV care services will be considered. This includes all observational studies (cross-sectional and cohort), interventional studies (single-arm intervention studies, randomised control trials, cluster randomised control trials, cross-over trials, and non-randomised control trials) and qualitative studies (interviews, surveys, focus groups, ethnography, phenomenology, grounded theory studies and qualitative process evaluations).
2. The target population for this review are AGYW living with HIV, which according to the WHO are females aged between 9 and 25 years of age. Therefore, only studies that include females within this age group living with HIV will be included.
3. Only studies published after 2006 will be considered, since the first commercially available prophylactic HPV vaccine was approved in this year.
4. This systematic review has global significance that aims to address universal challenges affecting cervical cancer research on a global scale and therefore there will be no geographical limits placed on search strategy.
5. We will not enforce any limitations on the language of publication. Instead, we will facilitate the translation of any potentially relevant publications into English to ensure their inclusion in the selection process and facilitate data extraction.

#### Exclusion criteria

1. Studies that exclusively focus on AGYW living without HIV or that do not clearly specify the age and HIV status of the study participants will be excluded as this review focuses primarily on AGYW living with HIV.
2. Studies that include females younger the 9 or older than 25 years of age as this does not align with the target population for this review.
3. Studies that evaluate the integration of cervical cancer prevention strategies into programs other than HIV care services will not be considered.
4. Studies published prior to 2006 will be excluded from this review as prophylactic HPV vaccines were not approved or commercially available at the time.
5. This review will exclude all other review studies, modelling studies, evidence synthesis studies, case studies, case series, case reports and conference proceedings.

#### Outcomes

1. A description of models used to integrate cervical cancer prevention strategies with existing HIV care programs will be identified and described.
2. The effectiveness of integrating cervical cancer prevention strategies into existing HIV care programs will be assessed in terms of the following outcomes:

2.1 The uptake of HPV vaccination, cervical cancer screening, precancerous treatment, and educational interventions by AGYW living with HIV following integration.
2.2 The knowledge, awareness, and willingness of AGYW living with HIV to utilise and adhere to cervical cancer prevention strategies following integration.

#### Search strategy

A search strategy will be developed by one of the authors (KG) together with an information specialist (SN). A comprehensive literature search will be performed to enable capturing of as many relevant articles as possible based on the inclusion and exclusion criteria outlined. The following online electronic databases will be searched: PubMed, Cochrane Central Library, EBSCO Host (Academic Search Premier, Africa-Wide Information, Cumulative Index to Nursing and Allied Health Literature [CINAHL], Health Source - Consumer Edition, Health Source: Nursing/Academic Edition, APA PsycArticles, APA PsycInfo), Web of Science, Scopus, and Google Scholar. Key words, medical subject headings (MeSH) and text words related to the themes; cervical cancer, HIV, prevention strategies and integration will be developed and then combined in the search strategy using Boolean operators, after which eligible articles will be identified. The search will be modified and applied to each electronic database (S1 Appendix. Database literature search strategy.). A standardized report template that will aid in keeping a record of all electronic databases searched, the search terms used and the total count of search results for each database will be developed and maintained. Reference lists of relevant studies will be searched for further articles in case they were missed during the primary searches. No geographical or language limitations will be applied.

#### Study selection

All publications identified from the electronic database searches will be downloaded into a reference manager program, Zotero [21], and imported into a web-based platform called Rayyan for deduplication and screening [22]. Two authors (KG and NB) will independently screen articles by title and abstract and then full text articles screened for inclusion against the eligibility criteria. Reasons for exclusion of full text studies that do not meet the inclusion criteria will be recorded and reported in a table. Where the same study, using the same sample and methods, has been presented in different reports, we will collate these reports so that each study (rather than each report) is the unit of interest in our review to avoid over representation of datasets in each study in the systematic review results. Oversight of the study selection process will be provided by a third author (EA-D). Any disagreements that arise between the two authors (KG and NB) at any stage of the study selection process will be resolved through discussion and will involve a third author (EA-D) if necessary. A PRISMA flow diagram will be used to present the selection process and results of the search [19].

### Data extraction and management

Data extraction will be performed using piloted extraction forms by two authors (KG and NB) to ensure consistency across included studies. Standardised data extraction forms will be designed using Microsoft excel file (Version 2402) and used to record extracted data from included publications. Individual forms will be created and used to record quantitative and qualitative data (S2 Appendix. Data extraction form) extracted from quantitative, qualitative, and mixed method studies. A pilot data extraction process with a draft extraction form will be performed on approximately 10 articles to determine if all relevant information is being captured.

The Cochrane Handbook will be used to provide guidance for the inclusion of cluster randomised control trials in this review [23].

Two authors (KG and NB) will extract the data, and a third author (EA-D) will crossLcheck the data to ensure that all relevant data has been extracted. Any disagreements between KG and NB will be resolved by discussion. A third author (EA-D) will be involved to resolve any outstanding disagreement as necessary.

### Dealing with missing data

Study authors will be contacted via email regarding any unreported data or to seek clarification on study methods. Should the data remain unavailable, the data at hand will be analysed and the significance of any missing data will be discussed among authors (KG, NB and EA-D). Authors will then deliberate on the most suitable method for dealing with the missing data as per guidelines outlined in the Cochrane Handbook [24].

### Methodological quality assessment

All included studies (quantitative, qualitative, and mixed methods) will be assessed for methodological quality independently and in duplicate by two authors (KG and NB). Any disagreements will be resolved by discussion or by involving a third author (EA-D) if necessary.

#### Quantitative studies

To assess randomised controlled trials (RCTs) included in this review we plan to utilise the Cochrane risk of bias tool (ROB-2) [25]. This tool evaluates selection, performance, detection, attrition, reporting and additional sources of bias allowing us to categorise each of the included studies as having low, moderate, or high risk of bias. A summary of the assessment of each study with the overall judgement will be recorded and tabulated. Non-randomised controlled trials (non RCTs) will be critically appraised using the Newcastle-Ottawa Scale (NOS) [26]. The NOS uses a star system for each included non RCT, which entails scoring stars based on a specific criterion. The overall quality of each study will be interpreted based on the total number of stars awarded. A comprehensive summary of all the assessments conducted, along with an overall judgement, will be recorded and tabulated.

#### Qualitative studies

The Critical Appraisal Skills Programme (CASP) quality assessment tool for qualitative studies will be applied to determine the rigour of qualitative methods used in included studies. The CASP tool will be used to examine the quality of a study in relation to 10 questions about research aims, appropriateness of methodology and design, recruitment strategy, data collection, researcher reflexivity, consideration of ethical issues, data analysis, statement of finding and the value of the research [27].

#### Mixed methods studies

The Mixed Methods Appraisal Tool (MMAT) will be applied to assess the risk of bias for mixed method studies [28]. The following criteria will be used to assess the risk of bias:

1. Is there an adequate rationale for using a mixed methods design to address the research question?
2. Are the different components of the study effectively integrated to answer the research question?
3. Are the outputs of the integration of qualitative and quantitative components adequately addressed?
4. Are divergences and inconsistencies between quantitative and qualitative results adequately addressed?
5. Do the different components of the study adhere to the quality criteria of each tradition of the methods involved?

### Unit of analysis

The unit of analysis for the quantitative component of this review will be primarily at the individual level. Studies that will be selected for inclusion in this review will assess the effectiveness of integrating cervical cancer prevention strategies into HIV care services in improving the uptake of these strategies by AGYW living with HIV globally. Key epidemiological measures such as changes in the incidence, mortality, or prevalence of cervical cancer as well as utilisation of cervical cancer prevention services and rates thereof will be analysed at an individual level.

The unit of analysis for the qualitative component will be thematic. Qualitative data from included studies will be analysed to identify common themes related to the effectiveness of integrating cervical cancer prevention strategies into HIV care services. Themes related to the knowledge, awareness, and willingness of AGYW living with HIV to utilise these services following integration will be analysed at an individual level.

### Data synthesis

#### Quantitative studies

The findings from the included quantitative studies will be narratively summarised and graphically illustrated [29]. Study results will be expressed as either numeric results or measures of association, with their associated variation and confidence intervals. The magnitude of heterogeneity between the included studies will then be assessed quantitatively using the I^2^ statistic [30]. I^2^ values will be interpreted as follows: 0 to 40% might not be important; 30 to 60% may represent moderate heterogeneity; 50 to 90% may represent substantial heterogeneity and 75 to 100% represents considerable heterogeneity [30]. The significance of heterogeneity will be determined by the p-value as outlined in the Cochrane Handbook [30]. For studies with moderate to significant heterogeneity, a random effects model will be used to obtain a pooled estimate of the outcome. If heterogeneity is between 0 to 40% a random effects model will also be used. In this instance, the random effects model will account for our lack of knowledge about why real, or apparent, intervention effects differ by considering the differences as if they were random [30].

Random effects models for meta-analysis will be performed using Review Manager software (RevMan 2020, V.5.4.1). If the heterogeneity detected is significantly high, a subgroup analysis will be performed to detect the possible sources [31]. A funnel plot will be used to assess for publication bias using R software (R Studio version 2023.03.0+386). Asymmetric distribution of the plot will indicate potential publication bias [32]. In addition, to statistically confirm whether the asymmetry is significant or not the Begg and Egger’s test will be performed using R software, where a p-value less than 0.05 indicates asymmetry and potential publication bias (R Studio version 2023.03.0+386) [32].

#### Qualitative studies

For the qualitative analysis thematic synthesis will be used to combine the findings of studies that describe the knowledge, awareness, and willingness of AGYW living with HIV to utilise and adhere to cervical cancer prevention strategies following integration [33]. The findings of included qualitative studies will be examined against the aims of this review, recurring patterns will be identified, and the qualitative data patterns will be interpreted by developing a coding framework. One article that closely answers the review objectives will be selected and used as a starting point to build a coding list. Two authors (KG and NB) will conduct “line-by-line” coding according to the content and meaning of the relevant findings of the article. The two authors (KG and NB) will subsequently discuss, with a third author (EA-D), the “free” codes, develop an initial coding list, and independently test this list on two additional articles to determine if and how well the concepts translate from one study to another. The three authors (KG, NB and EA-D) will subsequently discuss the codes emerging from the data and agree on a preliminary coding framework. The remaining studies will then be coded line-by-line using the agreed coding framework, adding new codes as necessary. Relevant findings, reported anywhere in the primary qualitative studies, will be coded. If new codes arise during the analysis process, a discussion among the three authors will be conducted (KG, NB and EA-D) and the coding list will be amended accordingly. Two authors (KG and NB) will revisit articles already coded to determine if the new codes apply or not. This process will continue until they have extracted data from all the included articles.

Data extraction will be verified by a third author (EA-D). Review findings will then be synthesised from the data that have been given the same codes across the studies. Findings will be shared with the third author (EA-D) to review. Finally, we will re-read the included studies to check that we have extracted all data relevant to the findings.

#### Combining quantitative and qualitative data

Following the synthesis of quantitative and qualitative data independently, they will be combined using the methods and suggestions provided in the Cochrane Handbook [34]. According to the Johanna Briggs guidelines on data synthesis and integration for mixed method reviews if the research question can be addressed by quantitative and qualitative research designs a convergent integrated approach will be followed [35]. However, if the review aims to explore various aspects or dimensions of a particular phenomenon of interest the convergent segregated approach will be followed [35].

### Sensitivity Analysis

To ensure the robustness and reliability of the findings of this review a sensitivity analysis will be conducted to assess the impact of the various methodological decisions on the results of this review and will be performed on both quantitative and qualitative components. The domains that will be considered include the quality of the included studies, sample size and the meta-analysis technique applied. If results remain consistent across the different analyses, the results can be considered robust as even with different decisions they remain the same/similar. If the results differ across sensitivity analyses, this is an indication that the result may need to be interpreted with caution [35].

### Assessment of quality of evidence

#### Quantitative studies

The quality of evidence for primary quantitative outcomes will be evaluated using the five Grading of Recommendations Assessment, Development and Evaluation (GRADE) criteria: risk of bias, consistency of effect, imprecision, indirectness, and publication bias [36]. To facilitate this process, we will utilize GRADEpro GDT software and include footnotes to elucidate any determinations made to downgrade the quality of evidence. We will use the study design of each included study as a determining factor of whether to upgrade (i.e. observational studies) or downgrade (i.e. RCTs) the quality of evidence.

An assessment of each outcome will be presented in a GRADE Evidence Profile. Two authors (KG and NB) will detail the number of studies, the number of participants, and the numerical result of the meta-analysis for each outcome. The effects of interventions on the outcomes included in the GRADE Evidence Profiles will be interpreted according to magnitude of effect and certainty of the evidence, using GRADE guidance on informative statements to combine size and certainty of an effect [36]. If meta-analysis is unsuitable or units of analysis are incomparable, results will be presented in a narrative ‘Summary of findings’ table format, with a recognition of the imprecision in evidence due to the absence of a quantitative effect measure [37]. This process will be reviewed by a third author (EA-D).

#### Qualitative studies

To evaluate the confidence in synthesized qualitative findings, the Grades of Recommendation, Assessment, Development, and Evaluation- Confidence in the Evidence from Qualitative Reviews (CERQual) approach will be used [38]. This approach encompasses four key domains: methodological limitations, relevance of contributing studies to the research question, coherence of study findings, and adequacy of data supporting the study findings. For each outcome, two authors (KG and NB) will consolidate the findings from these four domains and offer rationale to elucidate any determinations made to downgrade the quality of evidence. This process will be reviewed by a third author (EA-D).

## Discussion

This review aims to address a critical gap in the existing literature by proposing a comprehensive mixed-methods systematic review on the effectiveness of integrating cervical cancer prevention strategies into HIV care programs. The dual burden of cervical cancer and HIV presents a significant public health challenge, especially in resource-limited settings, where both conditions are prevalent. There is a need to develop effective and integrated approaches that comprehensively address these dual health concerns simultaneously among AGYW living with HIV. The proposed research design, a mixed-methods systematic review, is well-suited to capture both quantitative effectiveness measures and qualitative insights into the knowledge, awareness, and willingness of AGYW living with HIV. By incorporating vaccination, screening, pre-treatment and educational interventions into the assessment framework, this review aims to provide a holistic understanding of the potential benefits derived from the integration of prevention strategies. The strength of this review is the inclusion of current and up-to-date literature. Existing studies focus on isolated elements resulting in a fragmented understanding. The global search covering multiple databases enhances the robustness and reliability of this review.

## Conclusion

The anticipated outcomes of this systematic review could inform and improve implementation of current comprehensive cervical cancer prevention guidelines recommended by the WHO. Ultimately effective integration not only aligns with a holistic approach to women’s health but also contributes substantially towards improving health outcomes for AGYW living with HIV. Reducing cervical cancer incidence and mortality can make a meaningful impact on global public health, thus emphasizing the importance of this systematic review.

## Ethics

Ethics approval will not be required for this review as the work constitutes a secondary analysis of published research, which is already available in the public domain.

## Research to practice

The review team anticipates that the findings from this proposed systematic review will enhance equitable access to cervical cancer prevention services, thereby promoting the quality of life for AGYW living with HIV.

## Author contributions

**Conceptualization**: KG, NB and EA-D

**Methodology:** KG, SN, NB and EA-D

**Formal analysis:** KG, NB and EA-D

**Data curation:** KG and SN

**Writing (original draft and preparation):** KG

**Writing (review and editing):** SN, NB and EA-D

**Visualization:** KG, NB and EA-D

**Supervision:** NB and EA-D

**Project administration:** KG

## Supporting information

S1 Appendix. Database literature search strategy.

S1 Checklist. PRISMA-P 2015 checklist.

S2 Appendix. Data extraction form.

## Data Availability

No datasets will be generated or analysed for this systematic review protocol. All relevant data from this study will be made available upon study completion.

## Supporting information

S1 Checklist. PRISMA-P 2015 checklist.

S1 Appendix. Database literature search strategy. S2 Appendix. Data extraction form.

## References

1. Cervical cancer. [cited 26 May 2024]. Available: https://www.who.int/news-room/fact-sheets/detail/cervical-cancer

2. Okunade KS. Human papillomavirus and cervical cancer. J Obstet Gynaecol. 2020;40: 602–608. doi:10.1080/01443615.2019.1634030

3. Burmeister CA, Khan SF, Schäfer G, Mbatani N, Adams T, Moodley J, et al. Cervical cancer therapies: Current challenges and future perspectives. Tumour Virus Res. 2022;13: 200238. doi:10.1016/j.tvr.2022.200238

4. Williamson A-L. Recent Developments in Human Papillomavirus (HPV) Vaccinology. Viruses. 2023;15: 1440. doi:10.3390/v15071440

5. Stelzle D, Tanaka LF, Lee KK, Ibrahim Khalil A, Baussano I, Shah ASV, et al. Estimates of the global burden of cervical cancer associated with HIV. Lancet Glob Health. 2021;9: e161–e169. doi:10.1016/S2214-109X(20)30459-9

6. WHO guideline for screening and treatment of cervical pre-cancer lesions for cervical cancer prevention. Second edition. Geneva: World Health Organization; 2021.

7. Human papillomavirus (HPV). [cited 26 May 2024]. Available: https://www.who.int/teams/immunization-vaccines-and-biologicals/diseases/human-papillomavirus-vaccines-(HPV)

8. Zhang M, Sit JWH, Chan DNS, Akingbade O, Chan CWH. Educational Interventions to Promote Cervical Cancer Screening among Rural Populations: A Systematic Review. Int J Environ Res Public Health. 2022;19: 6874. doi:10.3390/ijerph19116874

9. Banerjee D, Mittal S, Mandal R, Basu P. Screening technologies for cervical cancer: Overview. CytoJournal. 2022;19: 23. doi:10.25259/CMAS_03_04_2021

10. Castle PE, Einstein MH, Sahasrabuddhe VV. Cervical cancer prevention and control in women living with human immunodeficiency virus. CA Cancer J Clin. 2021;71: 505–526. doi:10.3322/caac.21696

11. Singh D, Vignat J, Lorenzoni V, Eslahi M, Ginsburg O, Lauby-Secretan B, et al. Global estimates of incidence and mortality of cervical cancer in 2020: a baseline analysis of the WHO Global Cervical Cancer Elimination Initiative. Lancet Glob Health. 2023;11: e197–e206. doi:10.1016/S2214-109X(22)00501-0

12. Kakotkin VV, Semina EV, Zadorkina TG, Agapov MA. Prevention Strategies and Early Diagnosis of Cervical Cancer: Current State and Prospects. Diagnostics. 2023;13: 610. doi:10.3390/diagnostics13040610

13. Sigfrid L, Murphy G, Haldane V, Chuah FLH, Ong SE, Cervero-Liceras F, et al. Integrating cervical cancer with HIV healthcare services: A systematic review. Consolaro MEL, editor. PLOS ONE. 2017;12: e0181156. doi:10.1371/journal.pone.0181156

14. Mwanahamuntu MH, Sahasrabuddhe VV, Kapambwe S, Pfaendler KS, Chibwesha C, Mkumba G, et al. Advancing Cervical Cancer Prevention Initiatives in Resource-Constrained Settings: Insights from the Cervical Cancer Prevention Program in Zambia. PLoS Med. 2011;8: e1001032. doi:10.1371/journal.pmed.1001032

15. Huchko MJ, Bukusi EA, Cohen CR. Building capacity for cervical cancer screening in outpatient HIV clinics in the Nyanza province of western Kenya. Int J Gynecol Obstet. 2011;114: 106–110. doi:10.1016/j.ijgo.2011.02.009

16. Moon TD, SilvaLMatos C, Cordoso A, Baptista AJ, Sidat M, Vermund SH. Implementation of cervical cancer screening using visual inspection with acetic acid in rural Mozambique: successes and challenges using HIV care and treatment programme investments in Zambézia Province. J Int AIDS Soc. 2012;15: 17406. doi:10.7448/IAS.15.2.17406

17. Ramogola-Masire D, De Klerk R, Monare B, Ratshaa B, Friedman HM, Zetola NM. Cervical Cancer Prevention in HIV-Infected Women Using the “See and Treat” Approach in Botswana. JAIDS J Acquir Immune Defic Syndr. 2012;59: 308–313. doi:10.1097/QAI.0b013e3182426227

18. Lizarondo L, Stern C, Carrier J, Godfrey C, Rieger K, Salmond S, et al. Mixed methods systematic reviews. In: Aromataris E, Lockwood C, Porritt K, Pilla B, Jordan Z, editors. JBI Manual for Evidence Synthesis. JBI; 2024. doi:10.46658/JBIMES-24-07

19. Page MJ, McKenzie JE, Bossuyt PM, Boutron I, Hoffmann TC, Mulrow CD, et al. The PRISMA 2020 statement: an updated guideline for reporting systematic reviews. BMJ. 2021; n71. doi:10.1136/bmj.n71

20. PRISMA-P Group, Moher D, Shamseer L, Clarke M, Ghersi D, Liberati A, et al. Preferred reporting items for systematic review and meta-analysis protocols (PRISMA-P) 2015 statement. Syst Rev. 2015;4: 1. doi:10.1186/2046-4053-4-1

21. Zotero | Downloads. [cited 29 May 2024]. Available: https://www.zotero.org/download/

22. Ouzzani M, Hammady H, Fedorowicz Z, Elmagarmid A. Rayyan—a web and mobile app for systematic reviews. Syst Rev. 2016;5: 210. doi:10.1186/s13643-016-0384-4

23. Higgins JPT, Eldridge S, Li T. Chapter 23: Including variants on randomized trials. Cochrane Handbook for Systematic Reviews of Interventions version 64. Available: https://training.cochrane.org/handbook/current/chapter-23

24. Deeks JJ, Higgins JPT, Altman DG. Chapter 10.12: Analysing data and undertaking meta-analyses: Missing data. Cochrane Handbook for Systematic Reviews of Interventions version 6.4. Available: https://training.cochrane.org/handbook/current/chapter-10

25. Sterne JAC, Savović J, Page MJ, Elbers RG, Blencowe NS, Boutron I, et al. RoB 2: a revised tool for assessing risk of bias in randomised trials. BMJ. 2019; l4898. doi:10.1136/bmj.l4898

26. Wells G, Shea B, O’Connell D, Robertson J, Peterson J, Losos M, et al. The Newcastle-Ottawa Scale (NOS) for Assessing the Quality of Nonrandomized Studies in Meta-Analysis.

27. Long HA, French DP, Brooks JM. Optimising the value of the critical appraisal skills programme (CASP) tool for quality appraisal in qualitative evidence synthesis. Res Methods Med Health Sci. 2020;1: 31–42. doi:10.1177/2632084320947559

28. Hong QN, Fàbregues S, Bartlett G, Boardman F, Cargo M, Dagenais P, et al. The Mixed Methods Appraisal Tool (MMAT) version 2018 for information professionals and researchers. Educ Inf. 2018;34: 285–291. doi:10.3233/EFI-180221

29. Popay J, Roberts H, Sowden A, Petticrew M, Arai L, Rodgers M, et al. Guidance on the Conduct of Narrative Synthesis in Systematic Reviews.

30. Deeks JJ, Higgins JPT, Altman DG. Chapter 10.10: Analysing data and undertaking meta-analyses: Heterogeneity. Cochrane Handbook for Systematic Reviews of Interventions version 6.4. Available: https://training.cochrane.org/handbook/current/chapter-10

31. Deeks JJ, Higgins JPT, Altman DG. Chapter 10.11: Analysing data and undertaking meta-analyses: Investigating heterogeneity. Cochrane Handbook for Systematic Reviews of Interventions version 6.4. Available: https://training.cochrane.org/handbook/current/chapter-10

32. Tawfik GM, Dila KAS, Mohamed MYF, Tam DNH, Kien ND, Ahmed AM, et al. A step by step guide for conducting a systematic review and meta-analysis with simulation data. Trop Med Health. 2019;47: 46. doi:10.1186/s41182-019-0165-6

33. Thomas J, Harden A. Methods for the thematic synthesis of qualitative research in systematic reviews. BMC Med Res Methodol. 2008;8: 45. doi:10.1186/1471-2288-8-45

34. Flemming K, Booth A, Hannes K, Cargo M, Noyes J. Cochrane Qualitative and Implementation Methods Group guidance series—paper 6: reporting guidelines for qualitative, implementation, and process evaluation evidence syntheses. J Clin Epidemiol. 2018;97: 79–85. doi:10.1016/j.jclinepi.2017.10.022

35. Aromataris E, Munn Z. JBI manual for evidence synthesis. Adelaide, Australia: Joanna Briggs Institute; 2020.

36. Santesso N, Glenton C, Dahm P, Garner P, Akl EA, Alper B, et al. GRADE guidelines 26: informative statements to communicate the findings of systematic reviews of interventions. J Clin Epidemiol. 2020;119: 126–135. doi:10.1016/j.jclinepi.2019.10.014

37. Chan RJ, Webster J, Marquart L. Information interventions for orienting patients and their carers to cancer care facilities. Cochrane Consumers and Communication Group, editor. Cochrane Database Syst Rev. 2011 [cited 26 May 2024]. doi:10.1002/14651858.CD008273.pub2

38. Lewin S, Glenton C, Munthe-Kaas H, Carlsen B, Colvin CJ, Gülmezoglu M, et al. Using Qualitative Evidence in Decision Making for Health and Social Interventions: An Approach to Assess Confidence in Findings from Qualitative Evidence Syntheses (GRADE-CERQual). PLOS Med. 2015;12: e1001895. doi:10.1371/journal.pmed.1001895

