## Supplementary material for "Effectiveness of integrating cervical cancer prevention strategies into HIV care programmes: A mixed-methods systematic review protocol": S1 Appendix. Database literature search strategy.

Table 1: EBSCO Host (Academic Search Premier, Africa-Wide Information, Cumulative Index to Nursing and Allied Health Literature [CINAHL], Health Source - Consumer Edition, Health Source: Nursing/Academic Edition, APA PsycArticles, APA PsycInfo) literature search strategy.

| **Search** | **Query** | **Items found** |
| --- | --- | --- |
| #1 | Adolescent OR youth OR women OR “young women” OR female OR “young female” OR “young girl” OR “young female adolescent” | 8 063 144 |
| #2 | “human immunodeficiency virus” OR “human immuno deficiency virus” OR “human immune deficiency virus” OR hiv OR “hiv positive” OR “hiv infection*” OR “hiv aids” OR “living with hiv” OR “living with human immunodeficiency virus” | 661 634 |
| #3 | #1 AND #2 | 222 598 |
| #4 | “hpv vaccin*” OR “hpv immune*” OR “human papillomavirus vaccin*” OR “human papillomavirus immun*” OR “human papilloma virus vaccin*” OR “human papilloma virus immun*” OR “cervical cancer vaccin*” OR “cervical cancer immun*” | 21 812 |
| #5 | “cervical cancer screening” OR “early detection of cervical cancer” OR “hpv screening” OR “human papilloma virus screening” OR “human papillomavirus screening” OR “conventional cytology test*” OR “pap smear” OR “pap test” OR “pap smear based screening” OR “pap smear exam*” OR “pap smear screening*” OR “cervical smear” OR “vaginal smear” OR “papanicolaou smear” OR “papanicolaou smear screening” OR “papanicolaou smear test*” OR “cervical smear test” OR “papanicolaou test*” OR “cervical cytology” OR “cytology cervical smears” OR “papanicolaou stained smears” OR “liquid based cytology*” OR lbc OR “human papillomavirus dna test*” OR “human papilloma virus dna test*” OR “hpv dna test*” OR “human papillomavirus dna screening” OR “hpv dna screening” OR “human papilloma virus test*” OR “human papillomavirus test*” OR “hpv dna based tests” OR “hpv dna based testing” OR “human papilloma virus dna detection” OR “human papillomavirus dna detection” OR “visual inspection screening” OR “visual inspection of cervix*” OR “visual inspection with acetic*” OR “visual inspection with acetic acid via” OR “visual inspection with lugo*” OR “visual inspection with lugol s iodine vili” | 33 364 |
| #6 | “cervical cancer treatment” OR “hpv treatment” OR “human papillomavirus treatment” OR “hpv therapy” OR “cervical cancer therapy” OR “cervical cancer therapeutics” OR ablation OR “pre treatment” OR excision OR conization OR cryotherapy OR leep OR “loop electrosurgical excision*” OR “cone biopsy” OR laser* OR “cold coagulation” OR “large loop excision of transformation zone” OR lletz OR colposcopy | 863 901 |
| #7 | “cervical cancer education*” OR “hpv education” OR “human papillomavirus education” OR “cervical cancer knowledge*” OR “hpv knowledge*” OR “human papillomavirus knowledge” OR “human papilloma virus knowledge” OR “cervical cancer awareness” OR “hpv awareness” OR “human papillomavirus awareness” OR willingness | 113 888 |
| #8 | #4 OR #5 OR #6 OR #7 | 1 022 632 |
| #9 | “hiv care” OR “hiv services” OR “hiv clinics” OR “hiv centre*” OR “hiv healthcare*” OR clinic OR hospital OR “public health clinic*” OR “public health hospital*” OR “private healthcare facilities” OR “private healthcare facility” OR “public health facility” OR “public health facilities” OR “community based organisation*” OR “family planning clinic” OR “womens health clinic” OR “antenatal clinic” OR “maternity clinic” OR “outreach assessment clinics” OR “mobile clinics” OR “community based organization” OR “medical centre*” OR “primary healthcare facility” OR “primary healthcare clinic*” | 6 475 516 |
| #10 | #3 AND #8 AND #9 | 2 654 |
| Filters | 2006-2024, full text | 638 |
|  | Academic journals | 612 |
|  | Females | 145 |

Table 2: Cochrane Central Library literature search strategy.

| **Search** | **Query** | **Items found (TRIALS)** |
| --- | --- | --- |
| #1 | (Adolescent OR youth OR women OR “young women” OR female OR “young female” OR “young girl” OR “young female adolescent”):ti,ab,kw | 1 072 641 |
| #2 | (“human immunodeficiency virus” OR “human immuno deficiency virus” OR “human immune deficiency virus” OR hiv OR “hiv positive” OR hiv NEXT infection* OR “hiv aids” OR “living with hiv” OR “living with human immunodeficiency virus”):ti,ab,kw | 32 801 |
| #3 | #1 AND #2 | 20 158 |
| #4 | (hpv NEXT vaccin* OR hpv NEXT immune* OR human papillomavirus NEXT vaccin* OR human papillomavirus NEXT immun* OR human papilloma virus NEXT vaccin* OR human papilloma virus NEXT immun* OR cervical cancer NEXT vaccin* OR cervical cancer NEXT immun*):ti,ab,kw | 1 536 |
| #5 | (“cervical cancer screening” OR “early detection of cervical cancer” OR “hpv screening” OR “human papilloma virus screening” OR “human papillomavirus screening” OR conventional cytology NEXT test* OR “pap smear” OR “pap test” OR “pap smear based screening” OR pap smear NEXT exam* OR pap smear NEXT screening* OR “cervical smear” OR “vaginal smear” OR “papanicolaou smear” OR “papanicolaou smear screening” OR papanicolaou smear NEXT test* OR “cervical smear test” OR papanicolaou NEXT test* OR “cervical cytology” OR “cytology cervical smears” OR “papanicolaou stained smears” OR liquid based NEXT cytology* OR lbc OR human papillomavirus dna NEXT test* OR human papilloma virus dna NEXT test* OR hpv dna NEXT test* OR “human papillomavirus dna screening” OR “hpv dna screening” OR human papilloma virus NEXT test* OR human papillomavirus NEXT test* OR “hpv dna based tests” OR “hpv dna based testing” OR “human papilloma virus dna detection” OR “human papillomavirus dna detection” OR “visual inspection screening” OR visual inspection of NEXT cervix* OR visual inspection with NEXT acetic* OR “visual inspection with acetic acid via” OR visual inspection with NEXT lugo* OR “visual inspection with lugol s iodine vili”):ti,ab,kw | 2 969 |
| #6 | ("cervical cancer treatment" OR "cervical cancer therapy" OR "precancerous lesions treatment" OR "hpv treatment" OR "human papillomavirus treatment" OR "hpv therapy"):ti,ab,kw | 72 |
| #7 | (cervical cancer NEXT education* OR “hpv education” OR “human papillomavirus education” OR cervical cancer NEXT knowledge* OR hpv NEXT knowledge* OR “human papillomavirus knowledge” OR “human papilloma virus knowledge” OR “cervical cancer awareness” OR “hpv awareness” OR “human papillomavirus awareness” OR willingness):ti,ab,kw | 12 562 |
| #8 | #4 OR #5 OR #6 OR #7 | 16 740 |
| #9 | ("hiv care" OR "hiv services" OR "hiv clinics" OR "hiv centre" OR "hiv center" OR "hiv healthcare"):ti,ab,kw | 1 939 |
| #10 | #3 AND #8 AND #9 | 39 |
| Filters | 2006-2024 | 39 |

Table 3: PubMed literature search strategy.

| **Search** | **Query** | **Items found** |
| --- | --- | --- |
| #1 | Adolescent [Title/Abstract] OR youth [Title/Abstract] OR women [Title/Abstract] OR young women [Title/Abstract] OR female [Title/Abstract] OR young female [Title/Abstract] OR young girl [Title/Abstract] OR young female adolescent [Title/Abstract] | 2 252 942 |
| #2 | human immunodeficiency virus [Title/Abstract] OR human immuno deficiency virus [Title/Abstract] OR human immune deficiency virus [Title/Abstract] OR hiv [Title/Abstract] OR hiv positive [Title/Abstract] OR hiv infection* [Title/Abstract] OR hiv aids [Title/Abstract] OR living with hiv [Title/Abstract] OR living with human immunodeficiency virus [Title/Abstract] | 386 384 |
| #3 | #1 AND #2 | 62 688 |
| #4 | hpv vaccin* [Title/Abstract] OR hpv immune* [Title/Abstract] OR human papillomavirus vaccin* [Title/Abstract] OR human papillomavirus immun* [Title/Abstract] OR human papilloma virus vaccin* [Title/Abstract] OR human papilloma virus immun* [Title/Abstract] OR cervical cancer vaccin* [Title/Abstract] OR cervical cancer immun* [Title/Abstract] | 12 684 |
| #5 | cervical cancer screening [Title/Abstract] OR early detection of cervical cancer [Title/Abstract] OR hpv screening [Title/Abstract] OR human papilloma virus screening [Title/Abstract] OR human papillomavirus screening [Title/Abstract] OR conventional cytology test* [Title/Abstract] OR pap smear [Title/Abstract] OR pap test [Title/Abstract] OR pap smear based screening [Title/Abstract] OR pap smear exam* [Title/Abstract] OR pap smear screening* [Title/Abstract] OR cervical smear [Title/Abstract] OR vaginal smear [Title/Abstract] OR papanicolaou smear [Title/Abstract] OR papanicolaou smear screening [Title/Abstract] OR papanicolaou smear test* [Title/Abstract] OR cervical smear test [Title/Abstract] OR papanicolaou test* [Title/Abstract] OR cervical cytology [Title/Abstract] OR cytology cervical smears [Title/Abstract] OR papanicolaou stained smears [Title/Abstract] OR liquid based cytology* [Title/Abstract] OR lbc [Title/Abstract] OR human papillomavirus dna test* [Title/Abstract] OR human papilloma virus dna test* [Title/Abstract] OR hpv dna test* [Title/Abstract] OR human papillomavirus dna screening [Title/Abstract] OR hpv dna screening [Title/Abstract] OR human papilloma virus test* [Title/Abstract] OR human papillomavirus test* [Title/Abstract] OR hpv dna based tests [Title/Abstract] OR hpv dna based testing [Title/Abstract] OR human papilloma virus dna detection [Title/Abstract] OR human papillomavirus dna detection [Title/Abstract] OR visual inspection screening [Title/Abstract] OR visual inspection of cervix* [Title/Abstract] OR visual inspection with acetic* [Title/Abstract] OR visual inspection with acetic acid via [Title/Abstract] OR visual inspection with lugo* [Title/Abstract] OR visual inspection with lugol s iodine vili [Title/Abstract] | 25 559 |
| #6 | cervical cancer treatment [Title/Abstract] OR hpv treatment [Title/Abstract] OR human papillomavirus treatment [Title/Abstract] OR hpv therapy [Title/Abstract] OR cervical cancer therapy [Title/Abstract] OR cervical cancer therapeutics [Title/Abstract] OR ablation [Title/Abstract] OR pre treatment [Title/Abstract] OR excision [Title/Abstract] OR conization [Title/Abstract] OR cryotherapy [Title/Abstract] OR leep [Title/Abstract] OR loop electrosurgical excision* [Title/Abstract] OR cone biopsy [Title/Abstract] OR laser* [Title/Abstract] OR cold coagulation [Title/Abstract] OR large loop excision of transformation zone [Title/Abstract] OR lletz [Title/Abstract] OR colposcopy [Title/Abstract] | 645 512 |
| #7 | cervical cancer education* [Title/Abstract] OR hpv education [Title/Abstract] OR human papillomavirus education [Title/Abstract] OR cervical cancer knowledge* [Title/Abstract] OR hpv knowledge* [Title/Abstract] OR human papillomavirus knowledge [Title/Abstract] OR human papilloma virus knowledge [Title/Abstract] OR cervical cancer awareness [Title/Abstract] OR hpv awareness [Title/Abstract] OR human papillomavirus awareness [Title/Abstract] OR willingness [Title/Abstract] | 41 863 |
| #8 | #4 OR #5 OR #6 OR #7 | 717 571 |
| #9 | hiv care [Title/Abstract] OR hiv services [Title/Abstract] OR hiv clinics [Title/Abstract] OR hiv centre* [Title/Abstract] OR hiv healthcare* [Title/Abstract] OR clinic [Title/Abstract] OR hospital [Title/Abstract] OR public health clinic* [Title/Abstract] OR public health hospital* [Title/Abstract] OR private healthcare facilities [Title/Abstract] OR private healthcare facility [Title/Abstract] OR public health facility [Title/Abstract] OR public health facilities [Title/Abstract] OR community based organisation* [Title/Abstract] OR family planning clinic [Title/Abstract] OR womens health clinic [Title/Abstract] OR antenatal clinic [Title/Abstract] OR maternity clinic [Title/Abstract] OR outreach assessment clinics [Title/Abstract] OR mobile clinics [Title/Abstract] OR community based organization [Title/Abstract] OR medical centre* [Title/Abstract] OR primary healthcare facility [Title/Abstract] OR primary healthcare clinic* [Title/Abstract] | 1 603 438 |
| #10 | #3 AND #8 AND #9 | 710 |
| Filters | 2006 – 2024, Abstract, Full text, Female | 477 |

Table 4: Scopus literature search strategy.

| **Search** | **Query** | **Items found** |
| --- | --- | --- |
| #1 | TITLE-ABS-KEY ( adolescent OR youth OR women OR "young women" OR female OR "young female" OR "young girl" OR "young female adolescent" ) | 13 581 616 |
| #2 | TITLE-ABS-KEY ( "human immunodeficiency virus" OR "human immuno deficiency virus" OR "human immune deficiency virus" OR hiv OR "hiv positive" OR "hiv infection*" OR "hiv aids" OR "living with hiv" OR "living with human immunodeficiency virus" ) | 553 046 |
| #3 | #1 AND #2 | 225 287 |
| #4 | TITLE-ABS-KEY ( "hpv vaccin*" OR "hpv immune*" OR "human papillomavirus vaccin*" OR "human papillomavirus immun*" OR "human papilloma virus vaccin*" OR "human papilloma virus immun*" OR "cervical cancer vaccin*" OR "cervical cancer immun*" ) | 15 874 |
| #5 | TITLE-ABS-KEY ( "cervical cancer screening" OR "early detection of cervical cancer" OR "hpv screening" OR "human papilloma virus screening" OR "human papillomavirus screening" OR "conventional cytology test*" OR "pap smear" OR "pap test" OR "pap smear based screening" OR "pap smear exam*" OR "pap smear screening*" OR "cervical smear" OR "vaginal smear" OR "papanicolaou smear" OR "papanicolaou smear screening" OR "papanicolaou smear test*" OR "cervical smear test" OR "papanicolaou test*" OR "cervical cytology" OR "cytology cervical smears" OR "papanicolaou stained smears" OR "liquid based cytology*" OR lbc OR "human papillomavirus dna test*" OR "human papilloma virus dna test*" OR "hpv dna test*" OR "human papillomavirus dna screening" OR "hpv dna screening" OR "human papilloma virus test*" OR "human papillomavirus test*" OR "hpv dna based tests" OR "hpv dna based testing" OR "human papilloma virus dna detection" OR "human papillomavirus dna detection" OR "visual inspection screening" OR "visual inspection of cervix*" OR "visual inspection with acetic*" OR "visual inspection with acetic acid via" OR "visual inspection with lugo*" OR "visual inspection with lugol s iodine vili" ) | 53 463 |
| #6 | TITLE-ABS-KEY ( "cervical cancer treatment" OR "cervical cancer therapy" OR "precancerous lesions treatment" OR "hpv treatment" OR "human papillomavirus treatment" OR "hpv therapy" ) | 1 241 |
| #7 | TITLE-ABS-KEY ( "cervical cancer education*" OR "hpv education" OR "human papillomavirus education" OR "cervical cancer knowledge*" OR "hpv knowledge*" OR "human papillomavirus knowledge" OR "human papilloma virus knowledge" OR "cervical cancer awareness" OR "hpv awareness" OR "human papillomavirus awareness" OR willingness ) | 111 223 |
| #8 | #4 OR #5 OR #6 OR #7 | 177 408 |
| #9 | TITLE-ABS-KEY ( "hiv care" OR "hiv services" OR "hiv clinics" OR "hiv centre" OR "hiv center" OR "hiv healthcare" ) | 14 210 |
| #10 | #3 AND #8 AND #9 | 346 |
| #11 | 2006-2024 | 333 |
| #12 | Filters: Articles | 309 |

Table 5: Web of Science literature search strategy.

| **Search** | **Query** | **Items found** |
| --- | --- | --- |
| #1 | TS=(Adolescent OR youth OR women OR “young women” OR female OR “young female” OR “young girl” OR “young female adolescent”) | 3 579 021 |
| #2 | TS=(“human immunodeficiency virus” OR “human immuno deficiency virus” OR “human immune deficiency virus” OR hiv OR “hiv positive” OR “hiv infection*” OR “hiv aids” OR “living with hiv” OR “living with human immunodeficiency virus”) | 454 725 |
| #3 | #1 AND #2 | 83 388 |
| #4 | TS=(“hpv vaccin*” OR “hpv immune*” OR “human papillomavirus vaccin*” OR “human papillomavirus immun*” OR “human papilloma virus vaccin*” OR “human papilloma virus immun*” OR “cervical cancer vaccin*” OR “cervical cancer immun*” ) | 14 550 |
| #5 | TS=(“cervical cancer screening” OR “early detection of cervical cancer” OR “hpv screening” OR “human papilloma virus screening” OR “human papillomavirus screening” OR “conventional cytology test*” OR “pap smear” OR “pap test” OR “pap smear based screening” OR “pap smear exam*” OR “pap smear screening*” OR “cervical smear” OR “vaginal smear” OR “papanicolaou smear” OR “papanicolaou smear screening” OR “papanicolaou smear test*” OR “cervical smear test” OR “papanicolaou test*” OR “cervical cytology” OR “cytology cervical smears” OR “papanicolaou stained smears” OR “liquid based cytology*” OR lbc OR “human papillomavirus dna test*” OR “human papilloma virus dna test*” OR “hpv dna test*” OR “human papillomavirus dna screening” OR “hpv dna screening” OR “human papilloma virus test*” OR “human papillomavirus test*” OR “hpv dna based tests” OR “hpv dna based testing” OR “human papilloma virus dna detection” OR “human papillomavirus dna detection” OR “visual inspection screening” OR “visual inspection of cervix*” OR “visual inspection with acetic*” OR “visual inspection with acetic acid via” OR “visual inspection with lugo*” OR “visual inspection with lugol s iodine vili”) | 25 992 |
| #6 | TS=(“cervical cancer treatment” OR “cervical cancer therapy” OR “precancerous lesions treatment” OR “hpv treatment” OR “human papillomavirus treatment” OR “hpv therapy”) | 1 018 |
| #7 | TS=(“cervical cancer education*” OR “hpv education” OR “human papillomavirus education” OR “cervical cancer knowledge*” OR “hpv knowledge*” OR “human papillomavirus knowledge” OR “human papilloma virus knowledge” OR “cervical cancer awareness” OR “hpv awareness” OR “human papillomavirus awareness” OR willingness) | 87 657 |
| #8 | #4 OR #5 OR #6 OR #7 | 125 911 |
| #9 | TS=(“hiv care” OR “hiv services” OR “hiv centre” OR “hiv center” OR “hiv clinics” OR “hiv healthcare”) | 11 338 |
| #10 | #3 AND #8 AND #9 | 177 |
| Filters | 2006-2024 | 173 |
| Filters | Document types: "Article" and "Early Access" | 166 |

Table 6a: Google Scholar literature search strategy.

| **Search** | **Query** | **Items found** |
| --- | --- | --- |
| #1 | "adolescent girls and young women" AND "living with HIV" AND ("cervical cancer screening" OR "HPV vaccination" OR cervical cancer treatment") AND "HIV services" AND “integration” | 460 |
| Filters | 2006-2024 | 460 |

Table 6b: Google Scholar list of extracted articles.

| **Number** | **Google Scholar list of extracted articles** |
| --- | --- |
| 1 | Utilisation of cervical cancer screening among women living with HIV at Kenya’s national referral hospital. |
| 2 | Determinants of cervical cancer screening uptake among women of childbearing age in Mangochi district Malawi. |
| 3 | Modeling one-stop-shop integration of family planning and HIV services in Zimbabwe, 2021. |
| 4 | Sexual and reproductive health among adolescent girls and young women in Mombasa, Kenya. |
| 5 | Facilitators and barriers to cervical cancer screening among HIV-positive women in Ghana. |
| 6 | Implementation of cervical cancer prevention services for HIV-infected women in Zambia: measuring program effectiveness. |
