## Supplementary material for "Effectiveness of integrating cervical cancer prevention strategies into HIV care programmes: A mixed-methods systematic review protocol": S1 Checklist. PRISMA-P 2015 checklist.

**S1 Checklist. PRISMA-P (Preferred Reporting Items for Systematic review and Meta-Analysis Protocols) 2015 checklist.**

PRISMA-P Group, Moher D, Shamseer L, Clarke M, Ghersi D, Liberati A, et al. Preferred reporting items for systematic review and meta-analysis protocols (PRISMA-P) 2015 statement. Syst Rev. 2015;4: 1. doi:10.1186/2046-4053-4-1

| **Section/topic** | **#** | **Checklist item** | **Information reported** | | **Line number(s)** |
| --- | --- | --- | --- | --- | --- |
|  |  |  | **Yes** | **No** |  |
| **ADMINISTRATIVE INFORMATION** | | | | | |
| TITLE | | | | | |
| Identification | 1a | Identify the report as a protocol of a systematic review | x |  | 2-3 |
| Update | 1b | If the protocol is for an update of a previous systematic review, identify as such |  | x | NA |
| REGISTRATION | 2 | If registered, provide the name of the registry (e.g., PROSPERO) and registration number in the Abstract | x |  | 54-55 |
| AUTHORS | | | | | |
| Contact | 3a | Provide name, institutional affiliation, and e-mail address of all protocol authors; provide physical mailing address of corresponding author | x |  | 7-14 |
| Contributions | 3b | Describe contributions of protocol authors and identify the guarantor of the review | x |  | 583-592  16 |
| AMMENDMENTS | 4 | If the protocol represents an amendment of a previously completed or published protocol, identify as such and list changes; otherwise, state plan for documenting important protocol amendments |  | x | NA |
| SUPPORT | | | | | |
| Sources | 5a | Indicate sources of financial or other support for the review | x |  | 16 |
| Sponsor | 5b | Provide name for the review funder and/or sponsor |  | x | NA |
| Role of  sponsor/funder | 5c | Describe roles of funder(s), sponsor(s), and/or institution(s), if any, in developing the protocol |  | x | NA |
| **INTRODUCTION** | | | | | |
| RATIONALE | 6 | Describe the rationale for the review in the context of what is already known | x |  | 238-268 |
| OBJECTIVES | 7 | Provide an explicit statement of the question(s) the review will address with reference to participants, interventions, comparators, and outcomes (PICO) | x |  | 274-384 |
| **METHODS** | | | | | |
| ELIGIBILITY CRITERIA | 8 | Provide an explicit statement of the question(s) the review will address with reference to participants, interventions, comparators, and outcomes (PICO) | x |  | 299-334 |
| INFORMATION SOURCES | 9 | Describe all intended information sources (e.g., electronic databases, contact with study authors, trial registers, or other grey literature sources) with planned dates of coverage | x |  | 349-362 |
| SEARCH STRATEGY | 10 | Present draft of search strategy to be used for at least one electronic database, including planned limits, such that it could be repeated | x |  | S1 Appendix. |
| STUDY RECORDS | | | | | |
| Data management | 11a | Describe the mechanism(s) that will be used to manage records and data throughout the review | x |  | 379-393 |
| Selection process | 11b | State the process that will be used for selecting studies (e.g., two independent reviewers) through each phase of the review (i.e., screening, eligibility, and inclusion in meta-analysis) | x |  | 364-377 |
| Data collection  process | 11c | Describe planned method of extracting data from reports (e.g., piloting forms, done independently, in duplicate), any processes for obtaining and confirming data from investigators | x |  | 379-393 |
| DATA ITEMS | 12 | List and define all variables for which data will be sought (e.g., PICO items, funding sources), any pre-planned data assumptions and simplifications | x |  | 440-452 |
| OUTCOMES AND PRIORITISATION | 13 | List and define all outcomes for which data will be sought, including prioritization of main and additional outcomes, with rationale | x |  | 336-345 |
| RISK OF BIAS IN INDIVIDUAL STUDIES | 14 | Describe anticipated methods for assessing risk of bias of individual studies, including whether this will be done at the outcome or study level, or both; state how this information will be used in data synthesis | x |  | 402-438 |
| DATA | | | | | |
| SYNTHESIS | 15a | Describe criteria under which study data will be quantitatively synthesized | x |  | 455-476 |
|  | 15b | If data are appropriate for quantitative synthesis, describe planned summary measures, methods of handling data, and methods of combining data from studies, including any planned exploration of consistency (e.g., *I* ^2^, Kendall’s tau) | x |  | 455-476 |
|  | 15c | Describe any proposed additional analyses (e.g., sensitivity or subgroup analyses, meta-regression) | x |  | 510-518  470-471 |
|  | 15d | If quantitative synthesis is not appropriate, describe the type of summary planned | x |  | 456-457 |
| META-BIAS(ES) | 16 | Specify any planned assessment of meta-bias(es) (e.g., publication bias across studies, selective reporting within studies) | x |  | 471-476 |
| CONFIDENCE IN CUMULATIVE EVIDENCE | 17 | Describe how the strength of the body of evidence will be assessed (e.g., GRADE) | x |  | 520-546 |
