## Supplementary material for "Effectiveness of integrating cervical cancer prevention strategies into HIV care programmes: A mixed-methods systematic review protocol": S2 Appendix. Data extraction form.

|  | | STUDY CHARACTERISTICS | | | | | | | | CERVICAL CANCER PREVENTION STRATEGY |
| --- | --- | --- | --- | --- | --- | --- | --- | --- | --- | --- |
| Study  number | Title | | Author(s), Year published | Date of extraction | Country/  Region of study | Study design | Study aim | Sample size (before implementation of intervention) | Mean (sd)*/median (IQR)* age/age group of population |  |

Table 1a: Data extraction template for quantitative data extraction (Part 1)

*Note: sd= standard deviation; IQR= interquartile range

Table 1b: Data extraction template for quantitative data extraction (Part 2)

| METHOD AND DATA ANALYSIS | | | | | | | | | | | | | RELEVANT STUDY CONCLUSIONS | ANY ADDITIONAL COMMENTS |
| --- | --- | --- | --- | --- | --- | --- | --- | --- | --- | --- | --- | --- | --- | --- |
| Sample size (after implementation of intervention) | Data collection method | Outcome(s) assessed | | | | Unit(s) of measurement | | | | | | |  |  |
|  |  | Uptake of HPV vaccination | Uptake of cervical cancer screening | Uptake of precancerous treatment | Description of educational intervention (s) | Numeric result | | | Measure of association | | | |  |  |
|  |  |  |  |  |  | Scale | Result reported | Variance | Measure of association | Result reported | Upper statistical limit | Lower statistical limit |  |  |

| STUDY CHARACTERISTICS | | | | | | | | | | |
| --- | --- | --- | --- | --- | --- | --- | --- | --- | --- | --- |
| Study  number | Title | Author(s), Year published | Date of extraction | Country/  Region of study | Study design | Study aim | Study population | Mean age/age group of population | Number of participants | Cervical cancer prevention strategy |

| CERVICAL CANCER PREVENTION STRATEGY | METHOD AND DATA ANALYSIS | | | | CONCLUSION | COMMENTS |
| --- | --- | --- | --- | --- | --- | --- |
|  | Data collection method | Outcome(s) assessed and key findings (related to AGYW living with HIV to utilise and adhere to cervical cancer prevention strategies following integration into existing HIV care programs) | | |  |  |
|  |  | Knowledge | Awareness | Willingness |  |  |

Table 2a: Data extraction template for qualitative data extraction (Part 1)

Table 2b: Data extraction template for qualitative data extraction (Part 2)
